# Estimating Economic Burden and Quality of Life of Schistosomiasis in Madagascar and Burkina Faso

**DOI:** 10.64898/2026.08.12.26360326

**Authors:** Yongha Hwang, Jung-Seok Lee, Njariharinjakamampionona Rakotozandrindrainy, Lady Rosny Wandji Nana, Donghoon Kim, Chungman Chae, Gbènonminvo Enoch Cakpo, Ravomialisoa Razafimanantsoa, Lovasoa Derandrainy, Ndrainaharimira Rakotozandrindrainy, Eun Lyeong Park, Gabriel Nyirenda, Tabea Binger, Florian Marks, Raphaël Rakotozandrindrainy, Bassiahi Abdramane Soura

## Abstract

**Introduction:** Schistosomiasis is one of the most pervasive neglected tropical diseases in sub-Saharan Africa as an acute and chronic disease. It presents significant public health challenges in both Madagascar and Burkina Faso. Though schistosomiasis is a high prevalence disease, the understanding of its economic burden is limited.

**Methods:** A case control study was designed to compare the out-of-pocket costs and productivity loss associated with infection. A patient-specific survey questionnaire for Cost of Illness and Health-Related Quality of Life was developed and applied in Madagascar and Burkina Faso. Multiple interviews were conducted to capture the costs incurred during the entire period of illness.

**Results:** A total of 371 participants were enrolled in this study: 224 in Madagascar and 147 in Burkina Faso. The average economic burden for individuals confirmed to be negative for schistosomiasis was US$ 6.4 in Madagascar and US$ 29.8 in Burkina Faso. For mild cases, average costs were US$ 8.4 and US$ 13.5, respectively. Severe cases with complications due to schistosomiasis incurred substantially higher costs, averaging US$ 60.0 in Madagascar and US$ 1,272 in Burkina Faso. Median Health-Related Quality of Life was significantly lower among severe cases compared with controls and mild cases.

**Conclusions:** The cost of illness for schistosomiasis is significant in both countries. This study indicates the potential economic burden of disease in Madagascar and Burkina Faso, where the existing evidence on schistosomiasis is limited. Our study can help decision makers set health policy priorities.

## INTRODUCTION

In the past decade, there has been a notable increase in efforts to eliminate schistosomiasis, with the World Health Organization (WHO) setting 2030 as the goal for transmission interruption in endemic African countries (*1*). Schistosomiasis is an acute and chronic disease, affecting 251.4 million people and standing as one of the most pervasive neglected tropical diseases (NTDs) in sub-Saharan Africa (SSA) (*2*). This parasitic infection, primarily caused by *Schistosoma haematobium* and *S. Mansoni*, poses a substantial health burden throughout the countries (*1, 3*). The transmission of schistosomiasis occurs through contact with infested freshwater sources where parasitic larvae (cercariae) penetrate the skin (*2, 4, 5*). The release of eggs into the bloodstream can lead to severe clinical manifestations, including liver and kidney damage, infertility, cancer, and a range of other health complications (*6–8*).

In fact, schistosomiasis presents significant public health challenges, contributing to 200,000-540,000 deaths in SSA, where the disease is endemic (*9, 10*). Additionally, 90% of infected cases involve a substantial parasitic burden requiring preventive treatment (*6, 9, 11*). In Madagascar, schistosomiasis stands as a formidable health challenge, evident in its widespread prevalence, indicating it as endemic in 2016 (*9, 11*). An alarming estimate suggests that approximately 52.1% of Madagascar’s total population suffers from schistosomiasis, ranking it as the fifth-highest burdened nation globally(*9*). In Burkina Faso, a landlocked country situated in West Africa, the prevalence of schistosomiasis remains moderate on average but spikes drastically in specific regions such as Centre-Est, Est and Sahel, ranging between 20.94%-56.3% prevalence rate (*1, 2, 11, 12*). The country grapples with the concurrent burden of schistosomiasis, showing the endemic nature of public health challenges across its territories (*13*).

Schistosomiasis control strategies include mass treatment of at-risk groups, better water and sanitation, hygiene education, and snail control (*2*). WHO strategy of schistosomiasis control is mass administration of praziquantel targeted to at-risk population groups. However, praziquantel is effective against adult schistosome worms but has limited activity against immature larvae (*5*). It requires repeated treatment for curative parasite clearance. As a single drug administration does not prevent future infection, the cost-effectiveness of mass-treatment programs is compromised when reinfection is common.

Despite its widespread prevalence, there is a limited understanding of the economic burden of schistosomiasis, making it difficult to quantify its consequences across countries. Understanding the economic burden, particularly its cost-of-illness (COI), is important, as it enables us to develop and prioritize healthcare policies and interventions. Ultimately, this facilitates the allocation of healthcare resources in accordance with budgetary constraints, with the aim of achieving policy efficiency (*14, 15*).

A recent literature review revealed that, although the literature on control programs is extensive, only eight costing studies contained sufficient detail to analyze cost estimates (*16, 17*). Among these studies, only one estimated the total cost of the campaign implemented in Burkina Faso in 2004-2005, excluding out-of-pocket expenditures by patients. Additionally, no articles were found estimating the COI associated with Schistosomiasis in Madagascar (*18*). Notably, none of these studies include costs incurred by the individuals infected and their families; only the cost of control methods was considered. Given that schistosomiasis predominantly affects lower-income individuals (*19*), this gap in our understanding represents a significant limitation in assessing the true burden of the disease.

To address this knowledge gap, the current study focuses on estimating the cost-of-illness associated with schistosomiasis. By conducting a comprehensive analysis of health costs and productivity loss, this study aims to provide valuable insights into the economic burden of the disease on affected individuals and communities. This understanding is crucial for informing evidence-based healthcare policies and interventions, facilitating targeted resource allocation, and ultimately improving the efficiency of schistosomiasis control efforts. The objectives of this study are threefold: 1) To estimate the excess out-of-pocket health costs and productivity loss associated with schistosomiasis-infected individuals compared to uninfected individuals in Madagascar and Burkina Faso using a case-control design. 2) To estimate the cost-of-illness of individuals hospitalized due to schistosomiasis in Madagascar and Burkina Faso. 3) To measure the health-related quality of life (HRQoL) during infection in Madagascar and Burkina Faso.

## METHODS

### Surveillance and sites

The current study broadly consists of two activities: community-based enrolment (Activity 1), and hospital-based enrolment (Activity 2). For Activity 1, a case control study was designed to compare individuals who have been found to have schistosomiasis during the surveillance study to those who tested negative. On the other hand, Activity 2 concentrates on studying individuals with complicated or severe cases of schistosomiasis, primarily seeking treatment at specialized healthcare facilities.

To identify schistosomiasis, Point-of-Care Circulating Cathodic Antigen (POC-CCA) in urine was used for testing *S. mansoni* and filtration was also used for testing *S. haematobium* egg presence in urine. The patients were defined into three groups: 1) control case is laboratory-confirmed to be negative for *S. mansoni* and *S. haematobium*; 2) mild case is laboratory-confirmed positive for *S. mansoni* and *S. haematobium*; and 3) severe case is the patients that referred to the central health facility for complications due to schistosomiasis and laboratory-confirmed positive for schistosomiasis. Comparisons were made of their recent illnesses, healthcare seeking and ability to perform their usual activities in the periods before and after they were tested for schistosomiasis.

Study areas were selected from regions with high schistosomiasis prevalence in Madagascar and Burkina Faso (Supplementary Table 1). Control and mild cases were identified at the community level (Activity 1), and all participants were enrolled through the existing surveillance program called “Vaccine Against Schistosomiasis for Africa (VASA)”. In Madagascar, the study was conducted in two communes: Betafo and Soavina. The Betafo commune represents an urban population, located to the southwest of the capital, Antananarivo. It has a population of 33,250 and is comprised of 13 sub administrative units. The Soavina commune is located 10km away from Betafo and is a rural environment. There are seven sub-administrative units and a total population of 18,058. In Burkina Faso, the surveillance was conducted in four departments, namely Boromo, Manga, Houndé and Zorgho. These four departments have a combined population of 366,000. In these areas, the study was held in the villages of Vy, Bourzem, Pana and Zam respectively.

Severe cases were recruited at the health facility level (Activity 2). In Madagascar, the study team recruited participants in Ambrositra and Soavinandriana districts. There are approximately 21 Level-2 primary health centres (CSB IIs) which were used to recruit patients with complications. In Soavinandriana, only CSB IIs in commune Ampefy were included. These facilities were chosen as they were able to treat the complications of schistosomiasis, and the location of these facilities made the follow-up period feasible for the study team. In Burkina Faso, the study team conducted the study across the country. Patients who were presented at a health clinic and were suspected of complications due to schistosomiasis were referred to secondary and tertiary facilities in the country (Yalgado Hospital and two private clinics in particular). The study team asked doctors at these facilities to inform them if they had a patient with suspected schistosomiasis. The study team traveled to the facility to enroll the participants.

The standardized patient-level survey was implemented to collect the data across the two countries, and patients were interviewed up to six occasions in Madagascar and four occasions in Burkina Faso, depending on the seriousness and duration of their illness. All health-related costs were collected from three months prior to enrollment in the study until the last interview (up to 370 days).

### Study Design

The COI survey included three major cost components: direct medical cost (DMC), direct non-medical cost (DNMC), and indirect cost (IC). The DMC consists of consultation, medication, diagnostics fees, and all other costs which are directly related to the medical treatment for schistosomiasis. Patients were asked how much money they spent for medical services that they received. The DNMC includes any expenditure associated with food, lodging, and/or transportation for patients, as well as for the patients’ accompaniers. The IC takes account of the costs of productivity loss including wage loss, missing school days, and the costs associated with caregivers. To estimate productivity loss, their self-reported daily wage loss was asked of patients who had earnings. For students who did not have earnings, the government expenditure per primary student was used to convert their productivity loss (i.e., school absenteeism) into monetary value (*20, 21*). The government expenditure per primary student per day was estimated by dividing the annual expenditure by the number of school days, which was assumed to be 180 days per year. For those who had earnings but refused to report their wage, the minimum wage of each country was applied to calculate their productivity loss. If any patients passed away during the study period, productivity loss due to premature death was estimated based on the human capital approach (*22*): the age of death was subtracted from life expectancy obtained from the United Nations Population Division (*23*), and productivity years lost were then multiplied by the minimum wage, which was discounted at the rate of 3%. In addition to out-of-pocket or private cost estimation, information on non-out-of-pocket costs, such as public/private insurance coverage or governmental/health facility subsidies, was assessed for the severe cases at the health facility level.

The bootstrapping technique was applied to address uncertainty of the estimates and to take into account the skewed distribution of cost data. All estimates were presented with 95% confidence intervals (2.5th and 97.5th percentiles of the distribution) (*24*) and expressed in 2023 US$ using the official exchange rate from the World Bank (*25*).

In addition, HRQoL data were collected using the SF-20 survey form, a general health survey which is comprehensive and psychometrically sound, while still short enough for practical use (26). Because the SF-6D instrument developed by the University of Sheffield provides a preference-based framework for estimating health utilities, the SF-20 responses were mapped to the SF-6D (27). The SF-6D encompasses six dimensions: physical functioning, role limitation, social functioning, pain, mental health, and vitality. Owing to structural difference between the two instruments, the responses concerning role limitations in SF-20 items were collapsed into two categories (“limited” or “not limited”) to align with the corresponding SF-6D dimension. Health utility index was computed following the algorithm proposed by J. Brazier et al. (27), specifically using the mean model with an interaction effect and constant forced to unity (Model 10). If participants completed multiple surveys, the mean health utility index across all surveys was assigned to each individual. Because the coefficients for the three levels of role limitation (levels 2–4 in the SF-6D) in the original model fall within a narrow range (−0.050 to −0.055), a midpoint value of –0.0525 was applied uniformly to all role-limited responses from SF-20. The differences in median health utility index between each group were assessed. Depending on data distribution and variance homogeneity, either a two-way ANOVA (with Bonferroni correction for pairwise comparisons) or a Kruskal–Wallis test (with Dunn–Bonferroni post-hoc adjustment) was applied to assess group differences. Analyses were performed by using R and STATA version 17.

## RESULTS

### Basic Statistics

A total of 416 participants were enrolled. Of these, 6 in Madagascar and 39 in Burkina Faso were excluded due to loss to follow-up, as they could not be contacted or refused to continue. The final sample comprised in this study was 224 in Madagascar and 147 in Burkina Faso. The average age of the patients was highest in severe cases in Burkina Faso (56.1) and lowest in control cases in Madagascar (19.9). While there were more females among the participants in Madagascar, this was the opposite in Burkina Faso. The proportion of students among patients was higher in Madagascar than in Burkina Faso. The average monthly household income was estimated to be lower across all groups in Madagascar than in Burkina Faso (Table 1).

**Table 1.**
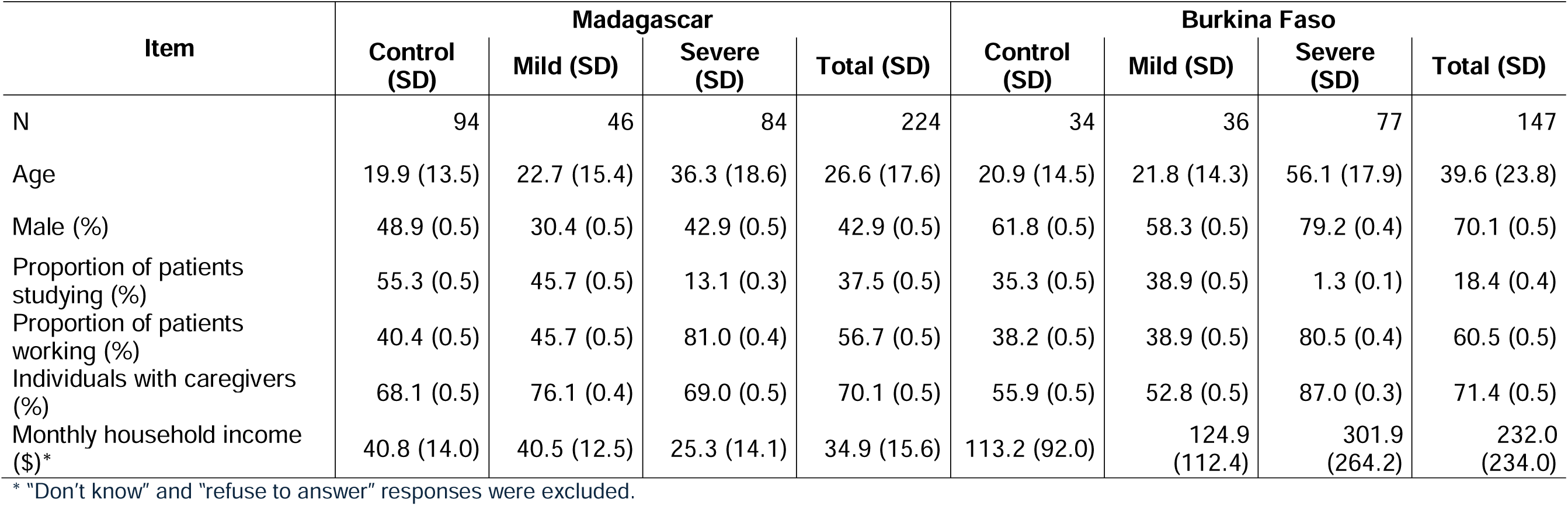
Descriptive statistics.

The age distribution of the control and mild cases are skewed to the right, meaning most of the patients were teenagers in both countries (Supplementary Figure 1). In contrast, the distribution of severe cases is relatively uniform across different age groups in Madagascar, while in Burkina Faso tended to be distributed among older age groups.

In Madagascar, the proportions of patients reporting cough, fever, and headache were high among control and mild cases (Supplementary Figure 2). In severe cases, the proportion reporting lethargy increased, while reporting fever decreased, with most patients reporting symptoms categorized as “other”. In Burkina Faso, fever and headache were also commonly reported in control and mild cases. In severe cases, the proportion reporting malnutrition increased, whereas most symptoms were reported less frequently compared with control and mild cases, reflecting a similar shift in symptom reporting. Among severe patients in both Madagascar and Burkina Faso, “other” symptoms were dominantly reported, including abdominal pain, ascites, urinary symptoms, dyspnea etc.

The health facilities visited by patients in each group are shown (Supplementary Figure 3), the percentage visiting a private and public facility was similar across all groups in Madagascar. In addition, the percent of visiting a pharmacy was the third highest in the control and mild groups. On the other hand, in Burkina Faso, the percentage of public facility visits accounted for the majority in control and mild cases, whereas private facility visits accounted for the majority in severe cases.

As expected, the economic burden per severe infection was greater than that of control or mild case groups in both countries (Figure 1). The total cost per schistosomiasis infection for severe cases was estimated to be US$ 60.0 in Madagascar and US$ 1,272.4 in Burkina Faso, while the total cost for mild cases was estimated to be US$ 8.4 and US$ 13.5, respectively. For both countries, the DMC component accounted for the biggest burden, while the proportion for the DNMC was the lowest.

**Figure 1.**
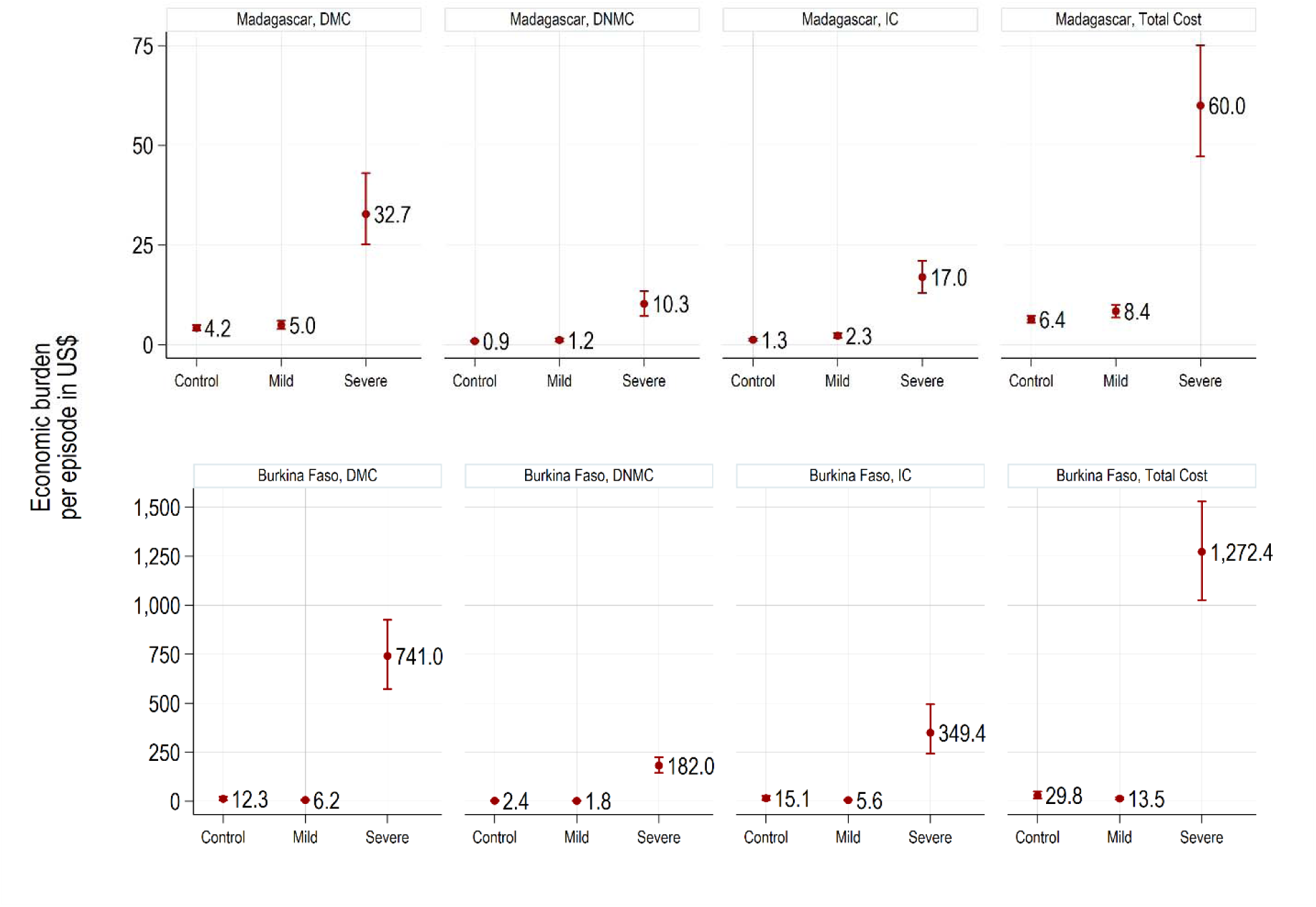
Economic burden of control, mild and severe cases by country.

The average economic burden of schistosomiasis was also estimated by age group (Figure 2). The total cost of schistosomiasis increased from younger cohorts (US$ 26.2) to older cohorts (US$ 76.7) in Madagascar severe cases. While the total cost among older cohorts was greater (US$1,427.7) than the cost among younger cohorts overall in Burkina Faso, the trend of the increase was not monotonous, and there was no statistically significant difference based on the Bonferroni Correction and Dunn-Bonferroni Test.

**Figure 2.**
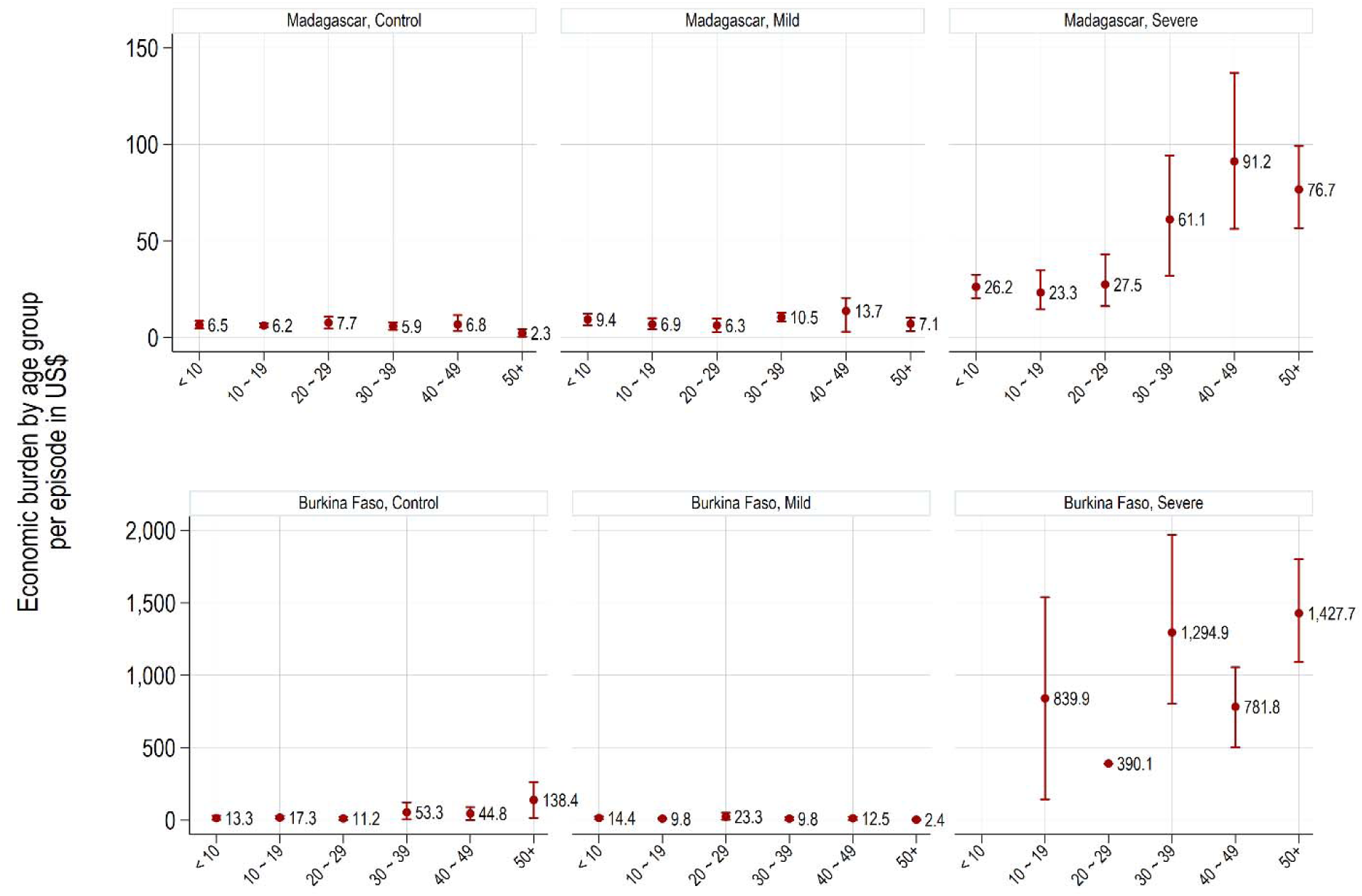
Economic burden of control, mild and severe cases by age groups.

In estimating the proportion of economic burden across cost components (Figure 3), in Madagascar, as cases changed from control to mild and severe, the proportion of DMC decreased from 66.0% to 54.6%, and the proportion of IC increased. In Burkina Faso, the proportion of DMC increased from 41.2% to 58.2%, and the proportion of IC decreased.

**Figure 3.**
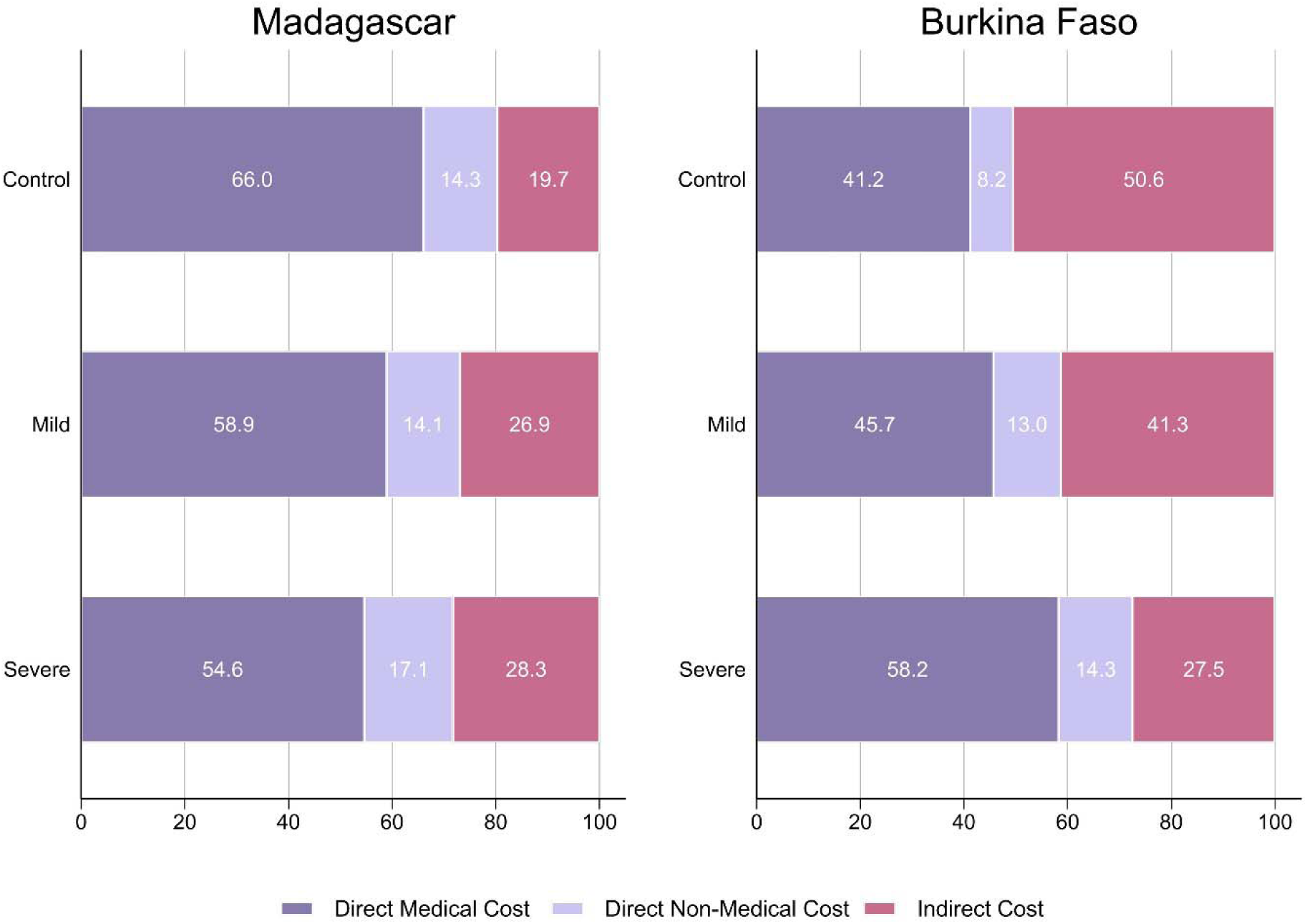
Portion of economic burden by cost components.

More details of the private costs of DMC for severe cases (Supplementary Table 2) show that in both countries, relatively higher costs were incurred at private hospitals/ clinics. However, it is important to note that the costs of private hospital/clinic in Burkina Faso were significantly higher.

While 32% of the total economic burden of severe schistosomiasis were covered by the public sector (insurance, government, health facility, etc.) in Madagascar, the public sector contribution (0.04%) was trivial in Burkina Faso (Supplementary Figure 4).

It should be noted that there were 28 deaths during the study period (10 in Madagascar, 18 in Burkina Faso). The average cost of productivity loss due to premature death was estimated to be US$ 7,726.0 for Madagascar, and US$ 5,153.0 for Burkina Faso.

### Health-related quality of life

The estimated HRQoL results are presented (Figure 4). In Madagascar, the median HRQoL was 0.841 (interquartile range [IQR] 0.803–0.877) in control cases, 0.814 (IQR 0.761–0.855) in mild cases, and 0.751 (IQR 0.700–0.796) in severe cases. The median HRQoL among severe cases was significantly lower than that in controls (p-value < 0.0001) and mild cases (p-value < 0.0001). In Burkina Faso, the median HRQoL was 0.806 (IQR 0.720–0.897) in control cases, 0.807 (IQR 0.724–0.865) in mild cases, and 0.741 (IQR 0.633–0.839) in severe cases. The median HRQoL among severe cases was significantly lower than that of controls (p = 0.015) and mild cases (p = 0.019), although with lower significance levels compared with Madagascar. In both countries, the difference in HRQoL between the control and mild case groups was not statistically significant.

**Figure 4.**
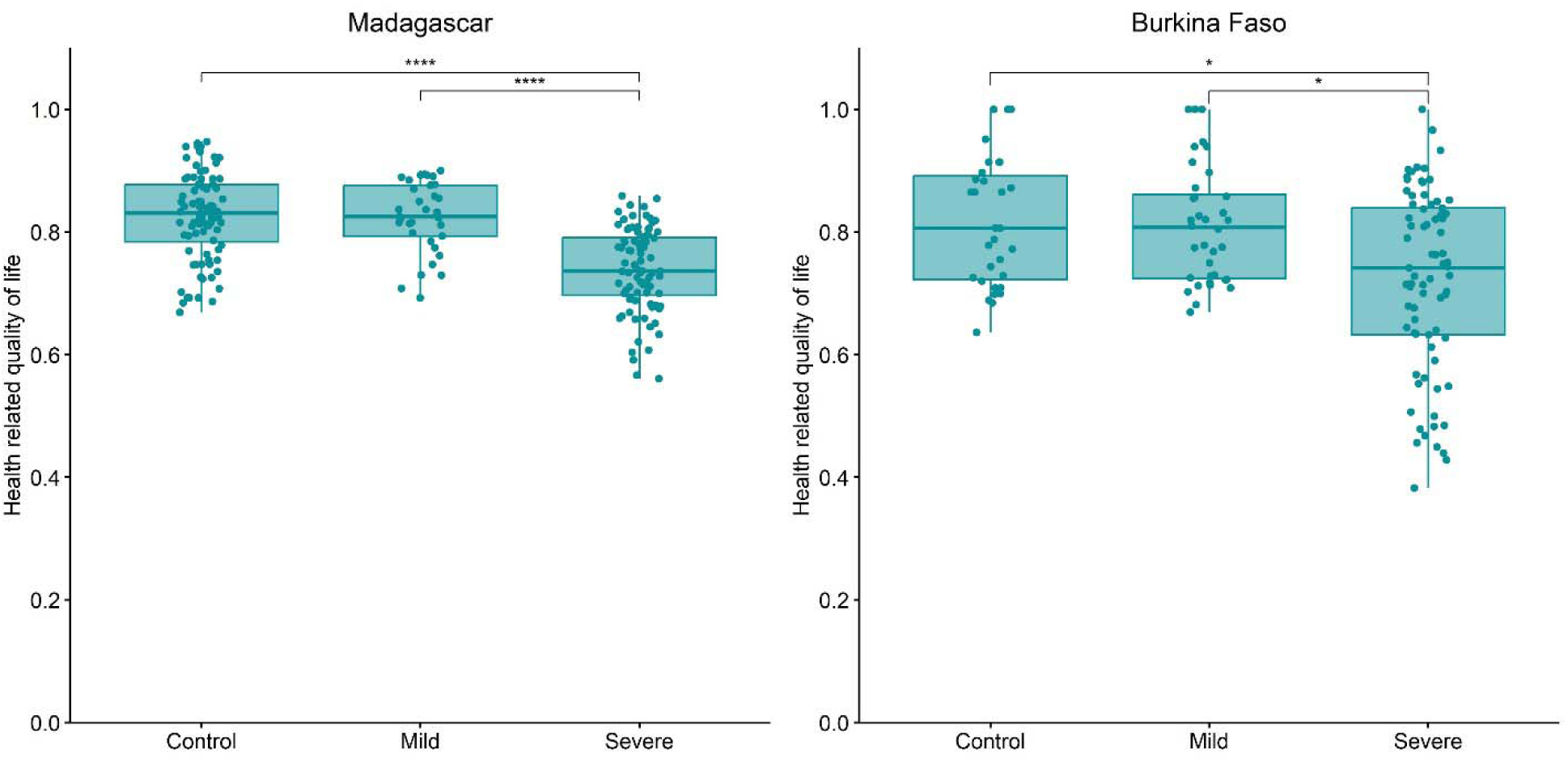
Health-related quality of life by country.

## DISCUSSION

The current study estimated the economic burden of schistosomiasis in a multi-country setting by implementing the standardized patient-level survey in Madagascar and Burkina Faso. In the context of schistosomiasis, there is little understanding of the effect of chronic infection on performance status or quality-of-life, as the disabling effects have not adequately been quantified. According to the systematic literature review on the existing economic burden of schistosomiasis (*17*), none of these studies include the costs incurred by the infected individuals and their families: only the cost of control methods is included.

Our study indicated that the economic burden of schistosomiasis is substantial in both countries. A recent review of the literature found that costs of schistosomiasis interventions range from US$ 0.06 to US$ 4.46 per person treated (*17*). While these estimates may not be directly comparable with our study, they are similar to the cost estimated for mild cases in our study.

However, with severe cases, the economic burden of schistosomiasis was much higher than mild cases: showing US$ 60.0 in Madagascar and US$ 1,272.4 in Burkina Faso. These findings highlight the importance of treating schistosomiasis before it becomes severe with complications. However, the substantial difference between the two countries may be attributable to variations in patient characteristics. For example, the average household income among severe-case patients was US$25.3 in Madagascar and US$301.9 in Burkina Faso—more than a tenfold difference—which may have been positively correlated with the economic burden. While considering only DMC for severe cases, they were US$ 32.7 and US$ 741. Madagascar had a much lower proportion of private facility use (15%) than Burkina Faso (68%), which may have contributed to the observed differences in treatment costs. They also show that patients in Burkina Faso had to bear most of economic burden, as very few patients were covered in the public sector. Lam HY et al. (28) estimated the financial cost of hospitalized schistosomiasis in the Philippines: the average cost of hospitalization for hepatic complications due to schistosomiasis was US$ 370.9, while uncomplicated schistosomiasis was US$ 66.1. Lam HY et al. also showed that the cost for hospitalization in private facilities averaged US$ 399.2, while in government facilities it was US$ 93.2 for neurological complications due to schistosomiasis.

In both Madagascar and Burkina Faso, there was no statistically significant difference in HRQoL between controls and mild cases. These findings suggest that mild schistosomiasis infection itself does not cause a measurable reduction in the HRQoL. However, severe cases resulting from heavy infection and associated complications significantly reduce the HRQoL. Studies assessing how severe schistosomiasis affect patients’ quality of life are limited, but several reported that the disease could lead to a decline in quality of life (29).

Some areas of uncertainty deserve attention. Firstly, the number of confirmed schistosomiasis infections was lower than expected in both countries. Furthermore, some patients left for other areas for several reasons, making follow-up interviews unfeasible, so the data could not be used for analysis. Secondly, the current study defined severe case patients as individuals with complications due to schistosomiasis, but this may differ from other studies which classifies intensity of schistosomiasis based on the estimated number of eggs (30). Thus, caution is required when comparing the data with other studies. Thirdly, the self-reported treatment costs were limited by participants’ ability to recall information from memory. The study team tried to minimize errors through multiple reviews, but there might still be some recall bias. Fourthly, the SF-6D has not been validated for national populations in African countries, except for one study that validated the SF-6D instrument for people living with HIV/AIDS in Kenya (31). Furthermore, most existing validations have used SF-6D index scores derived from SF-36 and SF-12 instruments. Because our study estimated HRQoL for schistosomiasis patients with SF-6D index scores derived from SF-20, differences in the structure and ordering of items in SF-20 compared with those in SF-36 and SF-12 may have influenced participants’ responses. Lastly, the current COI study was limited to the areas where the seroprevalence study was conducted. Only the severe cases for Burkina Faso were enrolled nationwide. Therefore, care should be taken when generalizing these estimates beyond the study communities.

Schistosomiasis, one of the most prevalent neglected tropical diseases, affects poverty, especially amongst the rural poor. Considering the importance of field-based economic evidence generation on schistosomiasis in the African region, the current study contributes to filling existing knowledge gaps in Madagascar and Burkina Faso. Future research is needed to have a better understanding of the disease and economic burden of schistosomiasis. For example, for decades, praziquantel has been the primary treatment for schistosomiasis. However, with its heavy reliance, praziquantel has raised concerns about schistosome resistance, particularly in areas with mass drug administration programs (32). Further studies comparing the economic burden of the infections which are susceptible to or resistance to praziquantel would contribute to understanding of schistosomiasis, as well as of the impact of future vaccines.

## DECLARATIONS

### Ethics approval and consent to participate

This study was conducted according to the guidelines laid down in the Declaration of Helsinki. The COI studies were approved by the Institutional Review Boards of the International Vaccine Institute, as well as by the ethical review committees of host country institutions: the Ethics Committee for Health Research in Burkina Faso, Comité d’Ethique de la recherche biomédicale auprès du Ministère de la Santé in Madagascar. All participants enrolled into the COI studies completed a written informed consent form. Children aged between 12 to 17 were provided with an explanation of the study. For children aged 5 to 12, their parents were asked to provide consent on the children’s behalf.

### Consent for publication

Not applicable

### Availability of data and materials

Aggregated data may be made available by the corresponding author upon reasonable request and subject to the conditions of the ethical approval.

### Authors’ contributions

Conceptualized, drafted the methods, and overall supervision J-SL; data analysis and drafted the rest of the manuscript YH; scientific support DK, CC, ELP, GN, TB, and FM; field support NR, LRWN, GEC, RR, LD, NR, RR, BAS.

## Acknowledgements

We would like to thank all members in the Vaccine Against Schistosomiasis for Africa (VASA) and local collaborators for their contributions and supports.

## Address for Correspondence

Name: Jung-Seok Lee

Address: International Vaccine Institute, SNU Research Park, 1 Gwanak-ro, Gwanak-gu, Seoul, 08826 Republic of Korea

Contact number: +8210 2840 3211

